# Supporting first FSH dosage for ovarian stimulation with Machine Learning

**DOI:** 10.1101/2022.02.10.22270790

**Authors:** N. Correa, J. Cerquides, J.L. Arcos, R. Vassena

## Abstract

**Research question:** Is it possible to identify accurately the optimal first dose of FSH in controlled ovarian stimulation (COS) by means of a machine learning (ML) model?

**Design:** Observational study (2011 to 2021) including first In Vitro Fertilization (IVF) cycles with own oocytes. 2713 patients from five private reproductive centers were included in the development phase of the model (2011 to 2019), and 774 in the validation phase (2020 to 2021). Predictor variables included: age, Body Mass Index (BMI), Antimullerian Hormone (AMH), Antral Follicle Count (AFC), and previous live births. Performance of the developed model was measured with a proposed score based on the number of MII retrieved and the dose received and/or recommended.

**Results:** The cycles included were from women 37.2±4.9 years old [18-45], with a BMI of 23.7±4.2, AMH of 2.4±2.3, AFC of 11.8±7.7; and an average number of MII obtained 7.2±5.3. The model reached a mean performance score of 0.87 (95% CI 0.86 to 0.88) in the development phase; this value was significantly better than the one for the doses prescribed by the clinicians for the same patients (0.83 [0.82, 0.84]; p-value= 2.44 e-10). The mean performance score of the model recommendations was 0.89 (95% CI 0.88 to 0.90) in the validation phase, also significantly better than clinicians (0.84 [0.82, 0.86]; p-value = 3.81 e-05). With these results the model was shown to surpass the performance of the standard practice.

**Conclusion(s):** The ML model developed could be deployed as a training and learning tool for new clinicians and serve as quality control for experienced ones; further, it could be used as second opinion, for instance by providing information in peer-to-peer case discussions.

**Key Message:** A Machine Learning model was trained to recommend first FSH doses for ovarian stimulation. When compared to clinicians the model developed had consistently better performance scores. The model could be used as a second opinion and as learning tool for new clinicians; to avoid as many non-optimal outcomes as possible.

## Introduction

Although significant strides have been made in the last 40 years, the mean pregnancy rate after an In Vitro Fertilization (IVF) cycle still hoovers around 30%, with a 20% chance of delivery (De Geyter et al., 2018). An important requisite to the success of an IVF cycle is the availability of a certain number of mature oocytes (MII); usually obtained after Controlled Ovarian Stimulation (COS).

COS represents therefore a key step for IVF success, as failing to ensure an optimal number of MII oocytes will likely hinder the rest of the procedure. As the number of MII oocytes retrieved increases, so does the chance of producing some embryos with high pregnancy potential (Drakopoulos et al., 2016; Esteves et al., 2019), but stimulating a patient too much leads to an increased risk of Ovarian Hyperstimulation Syndrome (OHSS). As such, a compromise must be reached to strive to retrieve a number inside of a range considered as optimal by literature, that does not increase chances of OHSS but maintains good pregnancy potential. The definition of an optimal range of oocytes we consider during this study goes from 10 to 15 oocytes and considers 4 to 9 oocytes as suboptimal (Polyzos & Sunkara, 2015). Anything outside these values is considered too much or too few. Whenever a patient falls outside the defined range, there is either the risk that the cycle is not successful or even cancelled, or the risk of OHSS, which also implies the need to freeze all the embryos in case they were generated, increasing the costs and delays in treatment.

Essential, then, to all COS protocols is the starting dose of exogenous Follicle Stimulating Hormone (FSH). This dose should be enough to recruit all FSH responsive follicles, but should not be any higher, to avoid unsafe effects such as OHSS or decreased oocyte quality. After approximately 8 days of stimulation, changing the FSH dose does not allow for a significant further recruitment of follicles (Fleming et al., 2006). In other words, if the starting dose of exogenous FSH is not adequate, little can be done to fix its effects on MII yield.

The choice of the FSH starting dose is mostly based on the patient’s characteristics such as age, Body Mass Index (BMI), or ovarian reserve and clinical characteristics such as past gravidity and parity. Sometimes, COS leads to unexpected and widely different results even among apparently similar patients, resulting in either too many or too few oocytes collected. Further, where the MII retrieved are in the expected number range, they may still be of insufficient quality to achieve success, as only 30-40% of microinjected oocytes develop to blastocyst (Maggiulli et al., 2020; Vaiarelli et al., 2020), and around 11% to an euploid blastocyst (Chamayou et al., 2017).

Clinicians use their knowledge and experience to prescribe a starting FSH dose to reach the appropriate range of follicular stimulation. So far, some Machine Learning (ML) models have been developed to encapsulate that medical experience reflected in historical data to try to automate that decision. Two separate nomograms based on patient age, Antimullerian Hormone (AMH) or Antral Follicle Count (AFC), and basal FSH levels have been developed for this task (Ebid et al., 2021; La Marca et al., 2012). One of them was tested prospectively (Allegra et al., 2017), reporting an increase in the number of patients with an optimal range of MII retrieved, and a decrease in patients with lower response in those using the nomogram. These two nomograms did not include patients older than 40 or those with irregular cycles, including Polycystic Ovary Syndrome (PCOS) patients. In a Randomized Controlled Trial (RCT) study for another model developed specifically for individualized dosage of FSH delta (Nyboe Andersen et al., 2017) no differences in pregnancy rate were observed.

Additionally, the CONSORT model, based on multivariate regression (Howles, Saunders, Alam, & Engrand, 2006) predicted overall lower starting doses compared to those prescribed by clinicians in normo-ovulatory patients (Naether et al., 2015; Pouly et al., 2015). CONSORT was also tested by RCT (Olivennes et al., 2015), showing that the model was able to reduce OHSS risk in patients while maintaining comparable pregnancy rates compared to the clinician chosen dose, in spite of a reduction in the number of retrieved oocytes.

The goal of this study is to develop and validate an ML based model designed to identify the optimal starting dose for all variants of FSH except delta (as it is not quantified in IU/ml), allowing to collect a number of MII as close as possible to 12 (a middle point in the optimal range considered in this study), for all types of patients.

## Materials and Methods

### Patient population and ethical approval

Data from a total of 2713 first IVF cycles registered in five private centers located in Spain and Italy, from 2011 to 2019, were used to develop the model. Natural cycles and cycles where gonadotrophins doses were not expressed in IU/ml were excluded. The inclusion of first cycles aimed to prevent bias due to unrecorded clinician knowledge (such as previous cycles’ FSH dosage and results). An additional 774 cycles from 2020 to 2021 were used for prospective validation of the model. Three categories of data were collected as variables. First, the input data, composed by age, BMI, proven fertility (Y/N) and reserve markers AMH and AFC. Second, the intervention, namely the first dose of FSH prescribed by the clinician. And third, result data expressed as number of metaphase II oocytes (MII) collected after stimulation (see Table I for details). During the whole study only cases with complete data on all the variables were included. Permission to conduct this study was requested and obtained from the Ethical Committee for Research of Eugin (approval code: 20/10/2020 ALGO2).

**Table I:** Patient characteristics in the two databases used in the study. Values are expressed as average and SD. Variables were compared using Mann-Whitney U test. For proportions a 2-sample z-test was conducted. A p-value of <0.05 was decided as significant.

|  | **Development**<br>**database (n=2713)** | **Validation**<br>**database (n=774)** | **p-value** |
| --- | --- | --- | --- |
| **Age (years)** | 37.9 ± 4.6 | 38.3 ± 4.4 | <0.05* |
| **AMH (ng/ml)** | 2.4 ± 2.3 | 2.2 ± 2.2 | <0.05* |
| **AFC (n)** | 11.1 ± 7.3 | 11.3 ± 8.5 | 0.7 |
| **BMI (kg/m<sup>2</sup>)** | 23.6 ± 4.2 | 23.2 ± 3.9 | <0.05* |
| **Number of MII**<br>**retrieved** | 6.8 ± 5.0 | 6.7 ± 6.5 | <0.05* |
| **Proven female**<br>**fertility** | 13% | 10.1% | 0.067 |

### Predictive model construction

In agreement with the current literature (Polyzos & Sunkara, 2015) we aimed at predicting the initial dose of FSH to achieve a number of MII as close as possible to 12, with the range 10-15 considered desirable, the range 4-9 suboptimal, and MII <4 or >15 not desirable. Given patient characteristics and limitations in the maximum dose of FSH administered, not every patient is considered able to reach the desired goal.

A predictive model was constructed to predict the patient’s capability to react to the first dose of FSH. This capability can be described by the slope of a simplified linear dose-response function. For any patient, during a natural cycle (0 UI/ml of exogenous FSH) the outcome in number of MII collected would stay mainly between 0 to 1. Given that in the database there are the results of a specific dose of FSH, the value of individual slopes is easily computable. To avoid negative slope values, it was assumed that all patients would get 0 MII if given 0 exogenous FSH.

The slope of a linear function is defined as follows:

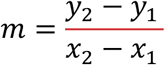

As the first data point (x_1_, y_1_) is set at the origin (0, 0), the slope value for every patient is computed by dividing the outcome or y_2_ (MII) by x_2_ (the first dose of FSH).

A Linear Regression algorithm was trained to predict the slope for every case (defined by its values at the start of the stimulation in age, BMI, AFC, AMH and proven fertility). Training was conducted on a random 80% of the development database. The remaining 20% was reserved for testing purposes. The training process was cross validated 5 times with 5 randomly selected training datasets, with their corresponding 5 test sets.

The final model used was composed by an intercept of 0.067, and coefficients for age, BMI, AFC, AMH an proven fertility of -0.00135, -0.00371, 0.00107, 0.00445 and -

0.00038 correspondingly.

### Dose recommendation by the model

For dose recommending purposes, the predicted slope for each test patient was used to compute the necessary FSH to get an outcome of 12 MII using the following linear function:

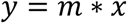

Where *y* is the number of MII, *m* the value of the slope, and *x* the FSH quantity.

As prescribing more than 300 UI/ml of FSH is described to give little to no advantages (Bastu et al., 2013; Harrison et al., 2001), recommended doses were capped at 300 UI/ml.

### Development of a performance score

We designed a score function to compare the model’s recommendations to the clinician prescriptions; given any FSH prescription with its resulting MII outcome, the score function assigns a score for a hypothetical recommended dose from -1 (the recommended dose was too low) to 1 (too high), being 0 the best possible value (the dose recommended was appropriate). FSH doses were categorized in 4 ordinal ranks (100 to 150, 151 to 200, 201 to 250, and 251 to 300) to create the score function.

The score function also allows assessing clinical prescriptions by setting the recommended dose equal to the clinician prescribed one. In doing so, the function evaluates how close the MII outcome is from the optimal range (10 to 15) and if there is any room for improvement dose-wise (see details in Appendix).

### Evaluation of performance of the model

Performance of model-based recommendations was evaluated using the proposed score in the reserved 20% for test of the development database and in the prospective validation database. In both cases, two scores were computed for each patient. One score for the dose prescribed by the clinician, and another score for the model recommended one. Absolute values of both scores were compared across all cases, in order to identify which group (clinical or model recommended) had more scores closer to 0, being here of no importance if the dose was too high or too low. The Wilcoxon signed-rank test was used for this purpose, as distribution of the scores was not normal. For an easier interpretation of the results, mean score values were expressed as 1 − |*score*|, where a value close to 1 is best. All computations were done using Scikit-learn 0.24 in Python 3.7.6.

## Results

### Predictive and recommendation performance

During the development phase of our model, the mean performance score for clinical doses was 0.83 (95% CI 0.82 to 0.84), and for model recommendations was 0.87 [0.86, 0.88] (p-value= 2.44 e-10).

During validation, the mean score for prescriptions was calculated to be 0.84 [0.82, 0.86], and for the model’s recommendations 0.89 [0.88, 0.90] (p-value = 3.81 e-05).

### Score and dosage analysis

To further understand the performance of the model and of the clinical prescriptions, we compared graphically the model, and the clinicians score distributions in the test set of the development database and in the validation one (Figure 1).

**Figure 1:**
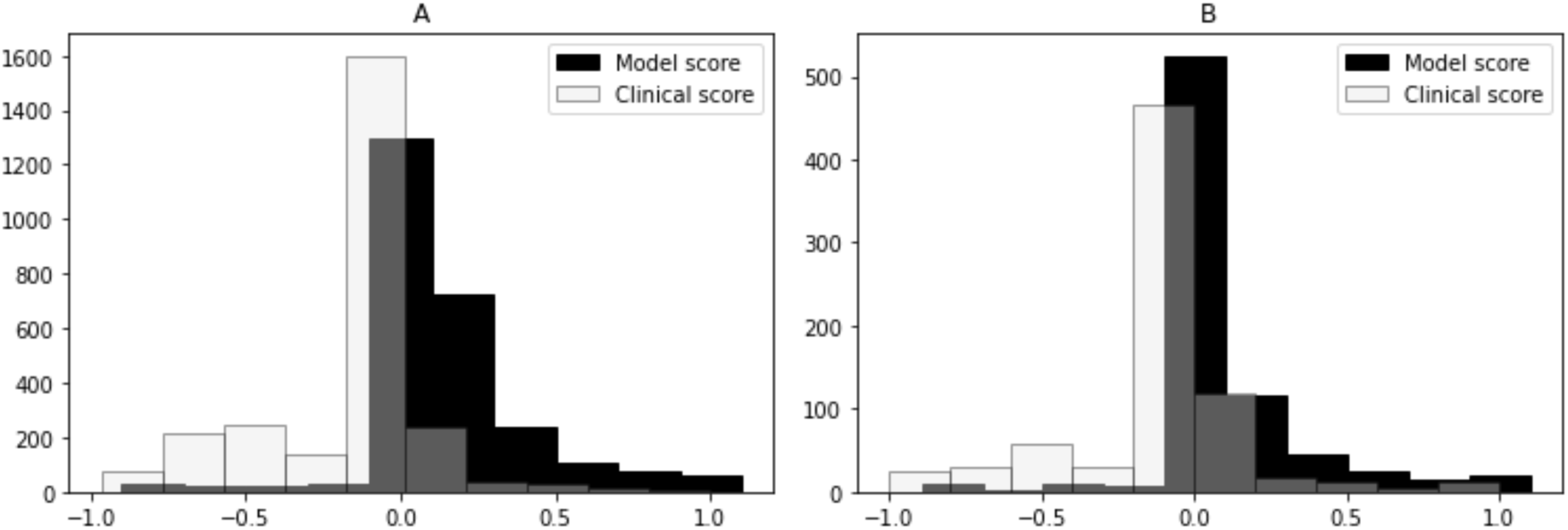
Performance during development and validation. Both clinical and model scores are plotted during development in panel A, and in panel B during validation. In black the model scores, and in light grey the clinical scores.

The model’s score approaches 0 (the best possible dose) more times than the clinician does and tends to suggest a dose higher than the one favored by the clinicians when not approaching 0. In 57.4% of cases in the test set and in 68.8% in the validation database the dose rank was not modified in relation to the clinician prescribed dose.

Further, we analyzed how the dosage was changed from clinician prescription to model recommendation, in relation to the real outcome in number of MII (Figure 2).

**Figure 2:**
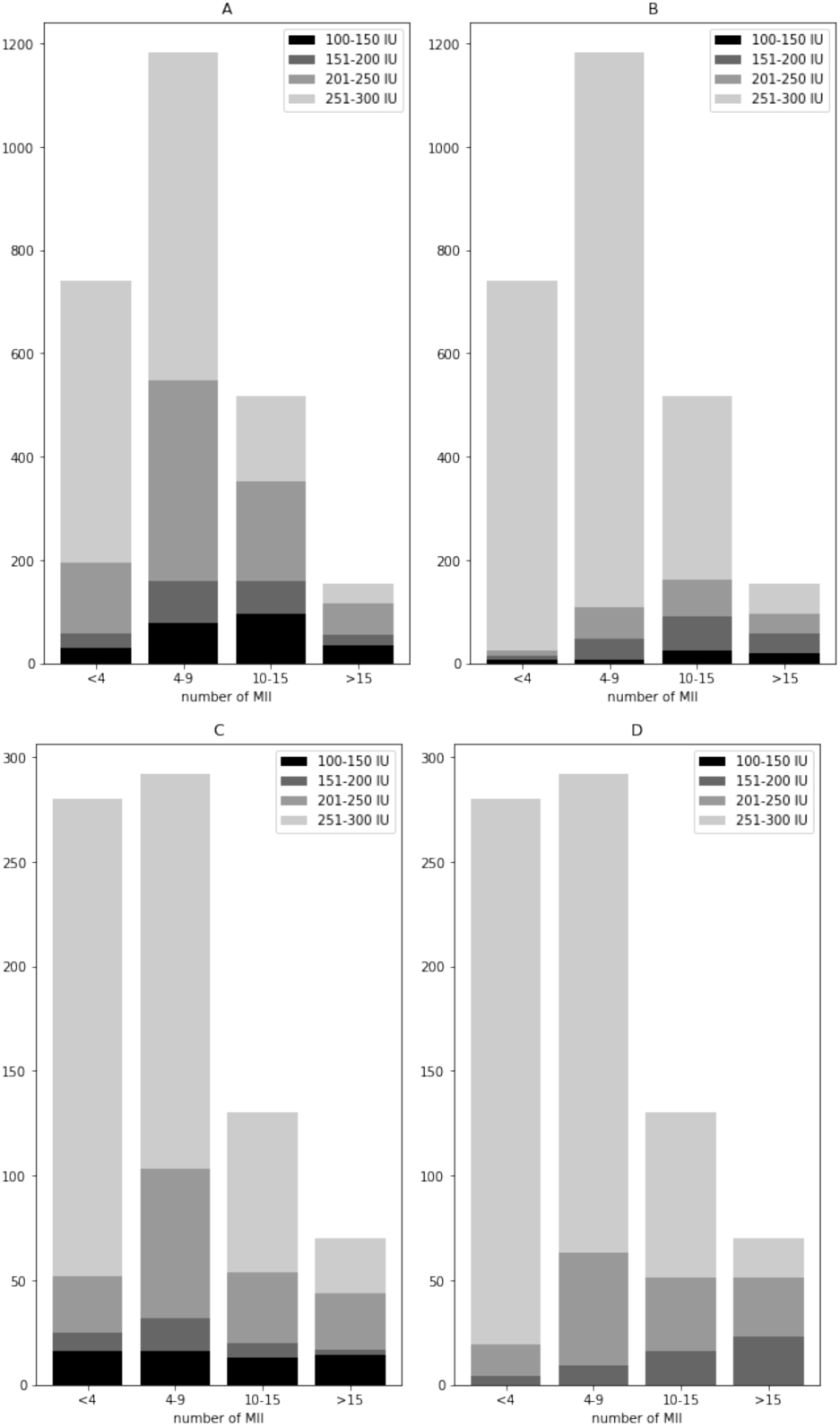
Dose ranks prescribed per bracket of MII retrieved. Panel A: by the clinicians during development. Panel B: by the model during development. Panel C: by the clinicians during validation. Panel D: by the model during validation. Dose ranks are represented lower to higher from black to light grey.

The model tends to increase the dose for patients with low and sub-optimal oocyte retrievals, but also increases dosage for some of the hyper-responders.

## Discussion

Current state of the art of FSH dosage models includes several recommender models that have provided very optimistic results. Yet, some of them have not been tested by an RCT study (Ebid et al., 2021) or the ones that do (Allegra et al., 2017; Nyboe Andersen et al., 2017; Olivennes et al., 2015) have not been developed to be used on all types of patients.

Including only normo-ovulatory patients (Howles et al., 2006), or younger than 40 patients with regular cycles (La Marca et al., 2012; Nyboe Andersen et al., 2017), is restricting this new personalization of the first FSH dose to a small subset of patients. And a subset, also, where this personalized dose finding is not as critical as with the patients excluded. The model presented in this study, improves state of the art in this aspect by including every type of patient to enhance results for all of them.

Including age, AFC, AMH, BMI and presence of previous successful pregnancies as variables in the core model has shown to allow for a good prediction of the dose-response function slope. This value has already been used as ovarian sensitivity (oocytes recovered per unit of starting FSH) in the development of an RCT tested nomogram (La Marca et al., 2012). Its use as objective variable of the core model mitigates the confounding effect that appears in any non-randomized treatment database, and that could lead a direct model (oocyte number as objective variable) to learn for example that higher doses, which are often prescribed for low-responders, lead to smaller oocyte yields. As in this setting this is always true (the treatment is tailored for the patient by the clinician), it is especially important to account for it. Removal of this effect, also allows for the extension of the recommender model for all types of patients, as they do not confuse the model, on the contrary, the core model learns that extreme patients (low-responders and hyper-responders) have extreme ovarian potential values.

As a separate contribution to an inclusive recommender model, we have developed a way to test in-silico whether the model would improve results versus the historical data, as a step preceding an RCT. To this end, the performance score was designed to encode and automate faithfully an expert clinical assessment of treatment-recommendation-outcome combinations. In other words, it enables us to assert if a recommended dose could fare better than the one already prescribed, given the real result in MII retrieved. In this way, it is possible to estimate reliably whether the model has the possibility to improve current clinical practice. With this information, the investment in a well-designed RCT can be made more confidently. Additionally, results of the in-silico performance of the model are more informative than the sole prediction scores of the core recommender model.

We show that, the scores of the model were consistently better than those of the clinical practice, both in the development database and the validation one. This is of interest as the model holds its value even though the population of the validation database is significantly older. Therefore, it means that the core model learnt the important aspects of the relationship between the patient’s characteristics and her ovarian potential or slope in the dose-response function. It is worth of note that the most significant predicted improvement was predicted for the patients whose oocyte yield was low or sub-optimal, where doses get increased on average. Upon implementation, the system’s recommendations may improve the average results and most probably avoid some cycle cancellations due to lack of embryos for transference.

Detailed analysis of the behavior of the model revealed its tendency, when wrong, to overdose some patients. This contrasts with clinical practice, where the tendency is to underdose when the prescription is not adequate. These instances of overestimations by the models correspond mainly to hyper-responder patient profiles, which are underrepresented on our databases. As such, the algorithm couldn’t learn appropriately due to the lack of a sufficient sample size. Importantly, while the model does tend to overdose these patients, it is of note that it still recommends the same or lower doses than the clinician in most of these cases, i.e. the clinician also tend to overdose. Nonetheless, we cannot dismiss the possibility that this could lead to a small increase in risk of OHSS. This contrasts with previous results in the literature, where RCT tested models reduced OHSS risk of incidence (Allegra et al., 2017; Nyboe Andersen et al., 2017; Olivennes et al., 2015). Secondary results of these studies, though, failed to show an increase either in retrieved oocytes or pregnancy results, with one reporting a reduction in oocyte yield (Olivennes et al., 2015). While OHSS risk must be taken very seriously, it is also true that nowadays it is manageable cycle-wise with proper prevention such as GnRH agonist trigger. All things considered, perhaps a manageable risk for a small portion of patients could be a fair trade-off to avoid a lack of embryos suitable for transference for other patients.

Further analysis of the instances where the model made a suboptimal suggestion led to another conclusion. The cases where the model gets negative error scores seem to coincide several times with also negative error scores for the clinician’s prescription. Looking into the specific cases where this happens gave us a profile of patients with very good markers and an unexplained low retrieval of oocytes. This could possibly be related to not diagnosed genetic polymorphisms in the FSHR or LHB genes (Lledo et al., 2014) that, obviously, nor the clinicians or the model could detect.

Despite the already described possible limitations of the system, it is encouraging that the preliminary results show in the vast majority of cases a similar or better performance score of the model’s recommendation versus the dose prescribed by the clinician.

## Conclusions

Clinicians prescribe the first FSH dose for each patient based in their characteristics, reserve markers and their own experience with similar cases. Although most of the time they prescribe the dose necessary for an optimal result, sometimes the outcome can unexpectedly vary and fall in suboptimal or extreme ranges. Our model could avoid most of these deviations by analyzing the patient’s profile and making a suggestion for the medical professional to assess.

Once tested and its performance confirmed via RCT, the ML model developed could be deployed as a training and learning tool for new clinicians and serve as quality control for experienced ones; further, it could be used as an second opinion, for instance by providing information in peer-to-peer case discussions.

## Supporting information

TRIPOD checklist

## Data Availability

All data produced in the present work are contained in the manuscript

## Authors’ roles

N.C. involved in study design, data collection and curation, analysis, model construction, interpretation and manuscript preparation. J.C. involved in analysis, model construction, interpretation and manuscript preparation. J.A. involved in analysis, model construction, interpretation and manuscript preparation. R.V. involved study supervision, expert knowledge, and manuscript preparation.

## Acknowledgements

We would like to thank dr. Maria Jesús López for kindly lending us her time and her expertise on Controlled Ovarian Stimulation protocols.

