## Appendix for "Supporting first FSH dosage for ovarian stimulation with Machine Learning"

### Construction of the performance score functions

To be able to assign a score for every possible combination of the 3 variables (MII number, prescribed dose, and recommended dose), first, values for key combinations were set and reviewed with expert clinicians. Specifically, for 5 specific values of MII (0, 6, 10, 15 and 25), a table was crafted with the scores (from -1 to 1) describing the effect of, given a real dose of FSH and its outcome, changing it for another dose (or maintaining it). For example, if the outcome was 0 MII and the dose prescribed 100-150 IU/ml, maintaining that dose as a recommendation would result in a score of -1, as the dose is clearly insufficient.

All tables and its graphic representations are available below.

Score values for cases with 0 MII retrieved:

| Real dose rank / Recommended dose rank | 100-150 IU | 175-200 IU | 225-250 IU | >250 IU |
|---|---|---|---|---|
| 100-150 IU | -1 | -1 | -1 | -1 |
| 175-200 IU | -0.2 | -0.90 | -0.95 | -0.95 |
| 225-250 IU | 0.1 | -0.01 | -0.7 | -0.9 |
| >250 IU | 0.15 | 0 | 0 | -0.01 |

Table II: Score values for every prescribed/recommended dose rank given that the result was 0 MII.

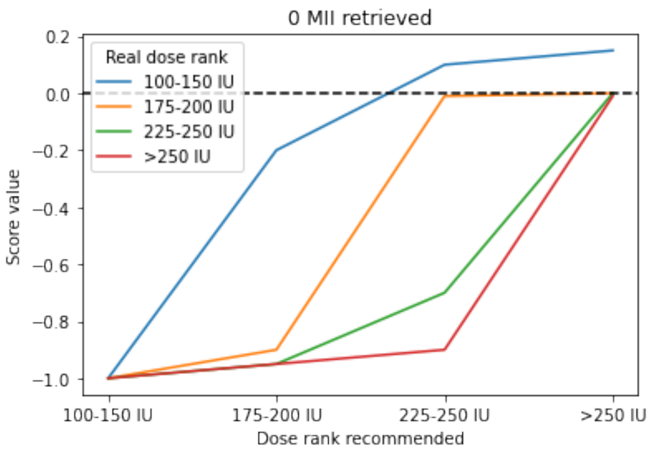


Figure 3: Linear representation of the scores of all combinations of prescription/recommended dose ranks given that the outcome was 0 MII retrieved.

<u>Scores values for cases with 6 MII retrieved:</u>

| **Real dose rank** / **Recommended dose rank** | **100-150 IU** | **175-200 IU** | **225-250 IU** | **>250 IU** |
|---|---|---|---|---|
| **100-150 IU** | -0.8 | -0.9 | -0.95 | -0.99 |
| **175-200 IU** | -0.05 | -0.60 | -0.85 | -0.9 |
| **225-250 IU** | 0.3 | 0 | -0.5 | -0.85 |
| **>250 IU** | 0.4 | 0.1 | 0 | -0.001 |

Table III: Score values for every prescribed/recommended dose rank given that the result was 6 MII.

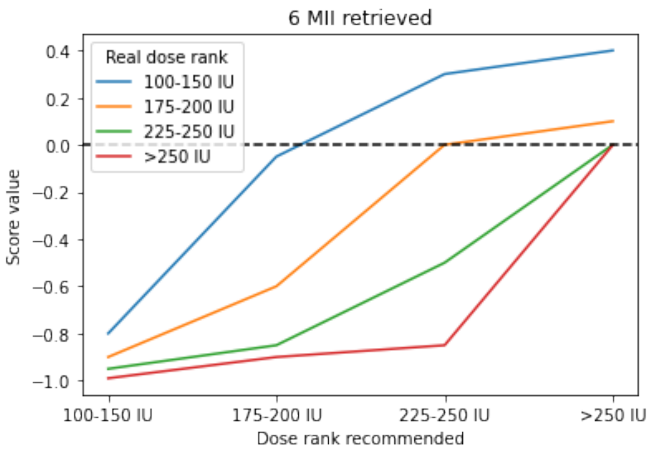


Figure 4: Linear representation of the scores of all combinations of prescription/recommended dose ranks given that the outcome was 6 MII retrieved.

<u>Score values for cases with 10 MII retrieved:</u>

| Real dose rank / Recommended dose rank | 100-150 IU | 175-200 IU | 225-250 IU | >250 IU |
|---|---|---|---|---|
| **100-150 IU** | -0.05 | -0.75 | -0.9 | -0.95 |
| **175-200 IU** | 0.1 | -0.01 | -0.75 | -0.8 |
| **225-250 IU** | 0.4 | 0.1 | -0.01 | -0.7 |
| **>250 IU** | 0.6 | 0.4 | 0.3 | 0 |

Table IV: Score values for every prescribed/recommended dose rank given that the result was 10 MII.

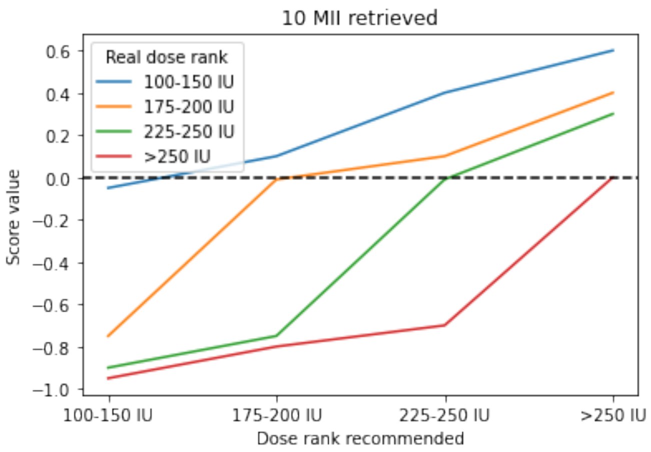


Figure 5: Linear representation of the scores of all combinations of prescription/recommended dose ranks given that the outcome was 10 MII retrieved.

Score values for cases with 15 MII retrieved:

| Real dose rank / Recommended dose rank | 100-150 IU | 175-200 IU | 225-250 IU | >250 IU |
|---|---|---|---|---|
| **100-150 IU** | 0.001 | -0.2 | -0.4 | -0.7 |
| **175-200 IU** | 0.2 | 0.01 | -0.15 | -0.6 |
| **225-250 IU** | 0.7 | 0.6 | 0.05 | -0.15 |
| **>250 IU** | 0.85 | 0.8 | 0.8 | 0.2 |

Table V: Score values for every prescribed/recommended dose rank given that the result was 15 MII.

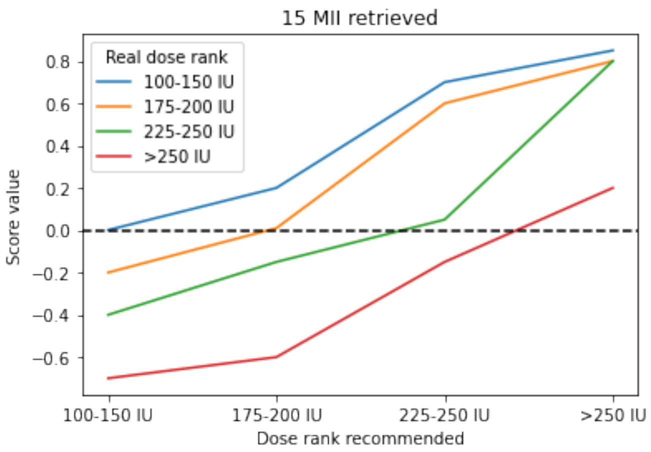


Figure 6: Linear representation of the scores of all combinations of prescription/recommended dose ranks given that the outcome was 15 MII retrieved.

<u>Score values for cases with 25 MII retrieved:</u>

| **Real dose rank** / **Recommended dose rank** | **100-150 IU** | **175-200 IU** | **225-250 IU** | **>250 IU** |
|---|---|---|---|---|
| **100-150 IU** | 0.01 | 0 | 0 | -0.1 |
| **175-200 IU** | 0.8 | 0.40 | 0 | -0.01 |
| **225-250 IU** | 0.9 | 0.85 | 0.8 | 0.1 |
| **>250 IU** | 1 | 1 | 1 | 1 |

Table VI: Score values for every prescribed/recommended dose rank given that the result was 25 MII.

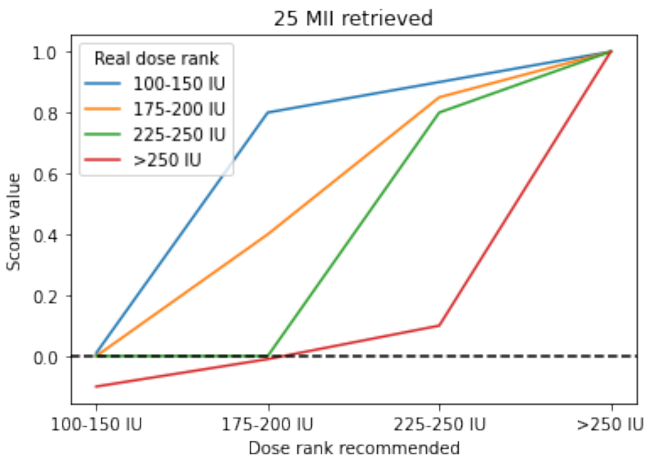


Figure 7: Linear representation of the scores of all combinations of prescription/recommended dose ranks given that the outcome was 25 MII retrieved.

For other combinations outside of the ones specified in the tables, linear regression was used. For every prescription/recommendation combination a function was stated using its score values across the 5 key values of MII as known points. When cases had values of MII recovered falling outside of the 5 values of MII stated, the corresponding function of prescription/recommendation was used to retrieve the resulting score.
